# Association of County-Level Socioeconomic and Political Characteristics with Engagement in Social Distancing for COVID-19

**DOI:** 10.1101/2020.04.06.20055632

**Authors:** Nolan M. Kavanagh, Rishi R. Goel, Atheendar S. Venkataramani

## Abstract

The U.S. is the epicenter of the coronavirus disease 2019 (COVID-19) pandemic. In response, governments have implemented measures to slow transmission through “social distancing.” However, the practice of social distancing may depend on prevailing socioeconomic conditions and beliefs. Using 15–17 million anonymized cell phone records, we find that lower per capita income and greater Republican orientation were associated with significantly reduced social distancing among U.S. counties. These associations persisted after adjusting for county-level sociodemographic and labor market characteristics as well as state fixed effects. These results may help policymakers and health professionals identify communities that are most vulnerable to transmission and direct resources and communications accordingly.

## Introduction

The U.S. is the epicenter of the coronavirus disease 2019 (COVID-19) pandemic.^1^ Modeling studies suggest that, without mitigation efforts, rising numbers of COVID-19 cases may overwhelm the health system’s capacity and result in millions of deaths.^2^ As a result, federal, state, and local governments have implemented a patchwork of public health measures to slow transmission through “social distancing.” These measures include orders to stay home and close schools and non-essential businesses. The effectiveness of social distancing policies requires significant engagement from communities. However, communities may face barriers to successfully adhere to these policies depending on prevailing socioeconomic conditions and political beliefs.^3^ This study examines the socioeconomic and political determinants of engagement in social distancing among U.S. counties.

## Methods

We obtained data on social distancing from Unacast, who measure county-level averages of distance traveled per person, based on 15–17 million anonymous cell phone users per day. Adherence to social distancing was conceptualized as changes in average distance traveled for March 19–28, 2020 (most recent data available), relative to matched days of a pre-COVID-19 reference week. The validity of this distancing measure is confirmed by the sharp decrease after declaration of a national emergency for COVID-19 (**Figure 1A**).

**Figure 1.**
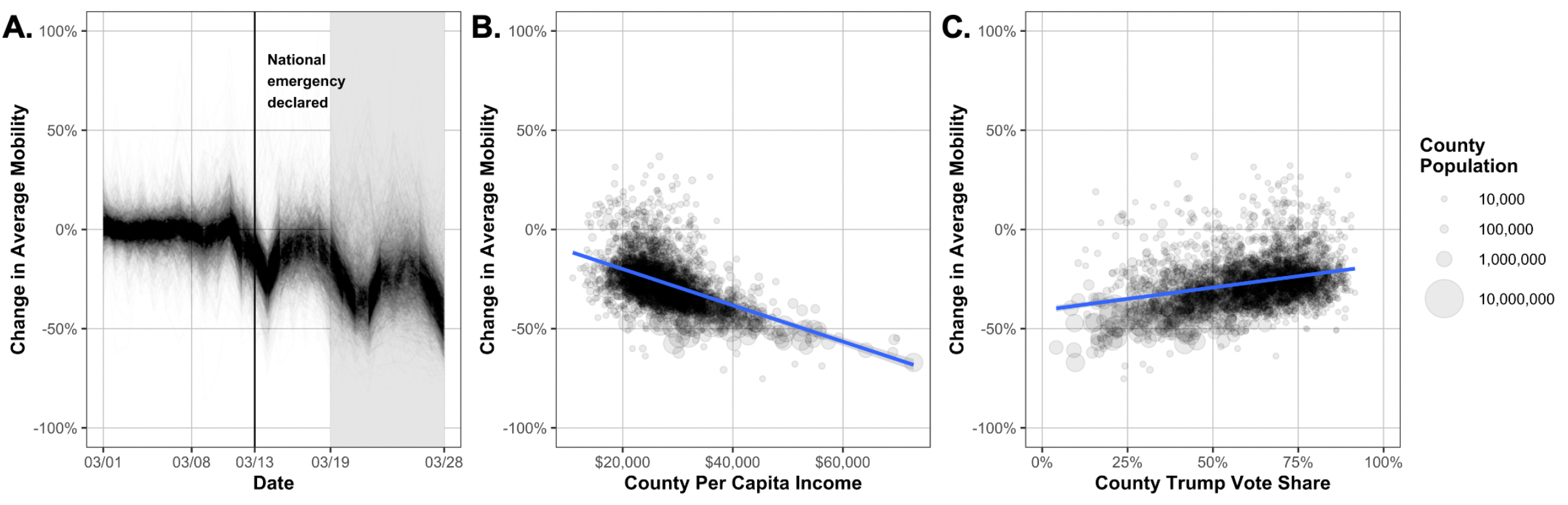
Association between Social Distancing and Income or Political Affiliation Among U.S. Counties. Notes: **(A.)** Social distancing among U.S. counties from March 1–28, relative to matched days of a pre-COVID-19 reference week. Mobility decreased by 27.3 percentage points in the median county during March 19–28. Associations between social distancing during March 19–28 and **(B.)** county per capita income or **(C.)** share of support for President Trump in 2016. Based on 3,037 counties, using anonymized cell phone records from Unacast and county-level data from the American Community Survey and MIT Election Data and Science Lab. Each line or dot represents one county; dots are sized according to population size. Fitted lines in **(B.)** and **(C.)** represent linear regressions with 95% confidence intervals, not adjusted for other county-level characteristics.

Data on the main exposures of socioeconomic status, operationalized by income per capita, and political preferences, operationalized by the 2016 county-level vote share for President Trump, were obtained from the American Community Survey (ACS, 5-year averages from 2014–2018) and MIT Election Data and Science Lab, respectively. Data on covariates (percentage male, Black, and Hispanic residents; age distribution; share of adults with college degrees; and shares of the workforce in industries most affected by COVID-19, i.e. retail, transportation, and health, educational, or social services) were also obtained from the ACS.

We estimated associations between the degree of social distancing and socioeconomic and political factors using cross-sectional ordinary least squares regressions, adjusting for sociodemographic covariates, state fixed effects (to account for state-level differences in timing and degree of COVID-19 mitigation measures), and rurality (from the 2010 Census).

## Results

In bivariate analyses, lower per capita income and a higher share of voters supporting President Trump were associated with reduced social distancing (**Figures 1B, 1C**). In multivariable models adjusting for sociodemographic and labor market characteristics as well as state fixed effects (**Table 1**), increasing per capita income from the bottom to top quartile (I.Q.R. = $7,218) resulted in a 1.4 percentage point increase in social distancing (95% C.I. = 0.2–2.5). An interquartile increase in support for Trump (I.Q.R. = 20.3%) resulted in a 4.1 percentage point decrease in social distancing (95% C.I. = 3.0–5.2). Higher shares of racial minorities and residents without college degrees were also associated with less distancing. There were no significant associations by population share of gender or age.

**Table 1.** Correlates of County-Level Change in Average Mobility from March 19–28, Relative to Matched Days of Pre-COVID-19 Reference Week.

| **Variable** | **Coefficient**<br>**($\beta$)** | **95% C.I.** | **P Value** |
| --- | --- | --- | --- |
| Per capita income | –1.36 | (–2.49, –0.23) | 0.018 |
| Share of Trump voters | 4.12 | (3.05, 5.19) | <0.001 |
| Percentage male | 0.33 | (–0.04, 0.70) | 0.079 |
| Percentage Black | 2.20 | (1.60, 2.81) | <0.001 |
| Percentage Hispanic | 1.09 | (0.69, 1.50) | <0.001 |
| Percentage with college degree | –2.76 | (–3.93, –1.60) | <0.001 |
| Employment in retail | 0.55 | (0.08, 1.02) | 0.023 |
| Employment in transportation | 1.23 | (0.75, 1.71) | <0.001 |
| Employment in health, education, social services | 0.89 | (0.32, 1.47) | 0.002 |
| Percentage rural | 0.83 | (–0.15, 1.81) | 0.10 |

Notes: Multivariable ordinary least squares regression for percentage-point change in average county mobility, given an interquartile change in given characteristics. Negative values indicate a greater degree of social distancing. The median county experienced a decrease in average mobility of 27.3 percentage points during March 19–28. Based on 3,037 counties, using data from Unacast, the American Community Survey, MIT Election Data and Science Lab, and Census. Model includes state fixed effects and percentage for each decade of life; there were no significant predictors among decades of life. 95% confidence intervals are provided.

## Discussion

Lower socioeconomic status and greater Republican political orientation were associated with reduced social distancing among U.S. counties. These results suggest that income-related barriers (e.g. the need to continue working) and political beliefs may lead to significant socioeconomic gradients in COVID-19, just as they do for other health conditions.^4-5^ Limitations of this study include the potential for omitted-variable and ecological biases due to aggregate, cross-sectional data. Social distancing may also be measured with error, given that the data do not sample all cell-phone users and do not reflect non-users.

Regardless, our findings underscore the heterogeneity of communities’ engagement in public health responses to COVID-19. These patterns may help policymakers and health professionals identify communities that are most vulnerable to transmission and direct resources and communications accordingly. Additional research as new data become available is necessary to correlate these findings with infection rates and mortality.

## Data Availability

All data from the American Community Survey, Census, and MIT Election Data and Science Lab are publicly available. Unacast provided us their data for research use; requests for access should be directed to them.

## Acknowledgments

We thank Unacast for providing us their social distancing dataset for research use.

